# MOKA: A pipeline for multi-omics bridged SNP-set kernel association test

**DOI:** 10.1101/2025.07.06.25330974

**Authors:** David Enoma, Dinghao Wang, Ariel Ghislain Kemogne Kamdoum, Rodrigo Ortega Polo, Quan Long, Jingni He

## Abstract

The explosion of genomic and multi-omics data has created a need for scalable, reproducible tools that integrate functional annotations into genome-wide association studies (GWAS). We introduce multi-omics data bridged Kernel Association test (MOKA) pipeline, a Snakemake-based workflow that automates SNP-set kernel-based association testing by incorporating multi-omics data, including gene expression, transcription factor binding, evolutionary conservation scores and neural network-derived features. This data-bridged architecture enhances variant prioritization and aggregation, improving statistical power in GWAS. MOKA supports population structure correction via spectral decomposition, parallel computation, and post-GWAS analyses, including visualization, Gene Ontology annotation, pathway enrichment, and validation. As a use case, we applied MOKA to a schizophrenia GWAS cohort, identified 89 Bonferroni-significant genes, with a 15.7% validation rate in disease-specific DisGeNET database and enrichment in pathways relevant to neuropsychiatric disease. MOKA provides a robust, scalable, and extensible framework for functional multi-omics integration in genetic studies. It is open-source and available at https://github.com/davidenoma/moka.

## 1. Introduction

Genome-wide association studies (GWAS) have been instrumental in identifying genetic variants associated with complex traits and diseases (Wu et al. 2010; Dehghan 2018). However, there is still a large proportion of unexplained heritability, particularly due to limitations in detecting the cumulative effects of causal variants (Manolio et al. 2009). Leveraging external and functional multi-omics data has shown promise in improving the power of GWAS by enabling more informed variant selection and aggregation (Wu et al. 2010).

Our previous research efforts have developed and applied the kernel-based association tests with the data-bridged architecture (Cao et al. 2021), which integrates external multi-omics data to guide variant prioritization and aggregation for downstream association mapping. This framework has been successfully applied to various data modalities, including gene expression (Cao et al. 2022a), brain imaging-derived phenotypes (He et al. 2024), transcription factor occupancy (He et al. 2022), and transcription factor binding-informed trans-variants (He et al. 2025). Additionally, a database resource has been developed to disseminate genome-wide analysis results (Cao et al. 2022b).

As the scale and diversity of genomic and multi-omics datasets continue to grow, there is an increasing demand for robust, scalable, and reproducible tools to streamline data processing and analysis. In bioinformatics, workflow management systems such as Snakemake (Koster and Rahmann 2018) have gained popularity due to their reproducibility, scalability, and ease of integration with high-performance computing environments. Despite the increasing complexity of the multi-omics data analysis (Cooper and Shendure 2011), there is a lack of automated pipelines tailored for kernel-based association tests, a class of statistical methods that model the joint effects of multiple variants in genetic studies.

To address this gap, we introduce the Multi-Omics Kernel-based Association (MOKA) pipeline, a fully automated analysis workflow built on the Snakemake workflow management system (Koster and Rahmann 2018). MOKA enables researchers and scientists to efficiently leverage the data-bridged kernel-based association tests in their GWAS datasets, boosting statistical power through the integration of diverse multi-omics data.

By leveraging Snakemake’s widespread adoption, extensive documentation, and active user community, MOKA ensures reproducibility, scalability, and ease of customization. This pipeline streamlines the entire analysis process, from multi-omics data integration and kernel-based association testing (with correction for population structure) to result visualization, biological annotation, disease database validation, Gene Ontology enrichment, pathway analysis. Additionally, this Snakemake pipeline also facilitates the harmonized use of tools from both Python and R, bridging distinct computational environments. Given the heterogeneity of data types and the complexity of parameter tuning in post-GWAS analysis, MOKA offers a robust and user-friendly solution for conducting comprehensive multi-omics association studies.

## 2. Methods

### 2.1. Design of MOKA

The MOKA pipeline is implemented using the Snakemake workflow management system (Koster and Rahmann 2018). Installation requires only a few simple steps, as outlined in the MOKA online documentation (https://github.com/davidenoma/moka).

### 2.2. Configuration, Data and software requirements

MOKA requires input genotype data in PLINK binary format (bed, bim, fam)(Purcell et al. 2007). Configuration is handled through a YAML file (config/config.yaml, see Table 1), which specifies input files, parameters, and auxiliary scripts. To optimize computational efficiency, MOKA supports chromosome-level parallelization by invoking GNU Parallel (Tange 2018) within a single Snakemake rule. Gene regions are defined as ±500 kb from gene boundaries based on the hg38 human genome reference (Mudge et al. 2025).

**Table 1.** Configuration table for MOKA. yaml format.

| Key | Purpose / Expected value |
| --- | --- |
| genotype_prefix | Basename of the PLINK trio (.bed/.bim/.fam). |
| weights_type | Short mnemonic for the bridge weights (e.g., phyloP, VAE-ED). |
| genotype_file_path | Folder containing the input PLINK files. |
| weights_file | Path to the CSV of variant weights. Header must include SNP_ID, Chromosome, Position, Weight. sample (weights/weights.csv) |
| gene_regions_file | CSV listing gene regions (Gene, Start, Stop, Chromosome) that define each SNP set. |
| disgenet_reference_file | Finally, disgenet_reference_file ("disease_database/{}.tsv") that points to a local cache of DisGeNET (Pinero et al. 2017) files. |
| plink_path | Absolute path to the PLINK executable (leave blank if PLINK is already on \$PATH). |
| is_binary | "TRUE" for case-control phenotypes, "FALSE" for quantitative traits. Guides downstream R scripts. |
| spectral_decorrelated | "TRUE" to apply GRM spectral decomposition (variance-component adjustment); "FALSE" to run the kernel test without this step. |

### 2.3. Multi-omics data bridge

Multi-omics data sources include user-defined functional genomic annotations at the nucleotide level, capturing biologically relevant features (Figure 1A), such as cis-regulatory variants, transcription factor binding (He et al. 2022; He et al. 2025), evolutionary conservation scores (Hubisz and Pollard 2014), gene expression changes (Li et al. 2024), imaging-derived weights, neural network-based (Li et al. 2023) approaches (Enoma et al. 2022), and others that promise to uncover disease-associated variants and genes (Cooper and Shendure 2011). All input weights are uniformly formatted across SNPs for integration into kernel models, with weight types customizable by the user.

**Figure 1.**
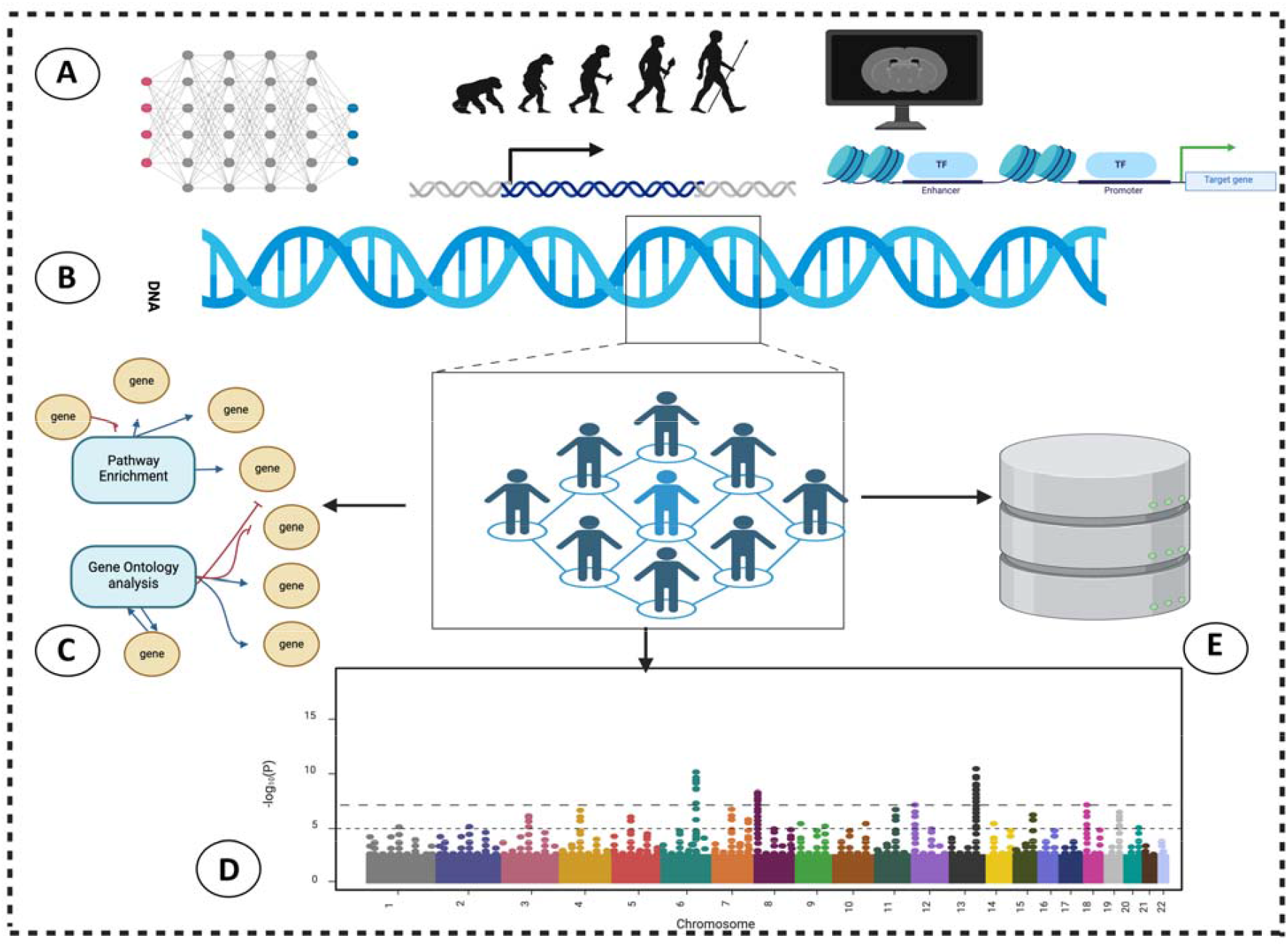
MOKA pipeline outlining the processes, including A. Diverse multi-omics data sources include variant-specific neural network weights, gene expression, conservation scores, brain image weights and regulatory element weights. B. multi-omics bridged association test on GWAS data. C. Gene Ontology and KEGG pathway enrichment analyses. D. Visualization of results in a Manhattan plot. E. DisGeNET disease database validation ratio of causal genes.

### 2.4. Kernel-based association testing with multi-omics weights

Within the input GWAS dataset, each SNP set or gene region is tested using a kernel-based method, which flexibly models epistatic and nonlinear effects of SNPs (Wu et al. 2010). For each individual, a weighted kernel is constructed using the SNPs and the corresponding data-bridged weights (Figure 1B).

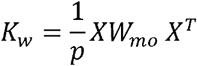

Where x is the genotype matrix for the SNPs in each gene region, *W*_*mo*_ is a diagonal matrix defined as 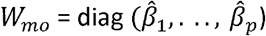, with *p* being the number of selected variants. The weights 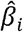 are derived from multi-omics data integration. This data-bridged weighted kernel (*K*_*W*_) is used in the association analysis implemented through a kernel-based test. The test statistic is defined as:

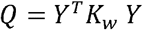

Where *Y* is the vector of phenotype values from the GWAS dataset, and *K*_*W*_ is the weighted kernel matrix defined above. Under the null hypothesis, *Q* follows a mixture of chi-squared distributions.

The Q-score test statistic evaluates whether the SNPs within the gene regions contribute to the observed phenotypic variation. The significance of the association for each gene region is assessed by calculating the p-value at a 0.05 threshold (prior to multiple testing correction), based on the null distribution of *Q*

### 2.5. Data transformation to control population structure

Uneven genetic relatedness will cause population structure, leading to inflated p-values (Kang et al. 2010). Linear Mixed Models are usually used for controlling this in single-SNP analysis (Lippert et al. 2011). However, here we are carrying out gene-based set test and aggregates SNPs within the gene region. To handle this problem, we use a transformation based on the decomposition of GRM. Previous work was done for both GWAS (Long et al. 2013) and gene expression analysis (Long et al. 2016) to decorrelated artifacts caused by uneven genetic relatedness using the same decompositions which we implement as a feature in the MOKA snakemake pipeline (https://github.com/davidenoma/moka).

Assuming the Linear Mixed model without fixed effect of a SNP takes form:

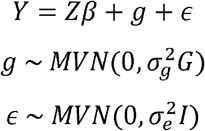

Where *Y* is the phenotype vector with *n* samples, *z* is the covariate matrix, and r is the vector of fixed effect sizes. *g* represents the random genetic effects, with 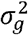 denoting the genetic variance and *G* being the genomic relationship matrix (GRM). ∈ denotes the residual effects, and 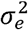 is the residual variance. Then, the variance of *Y* is given by 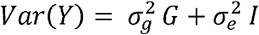 and the unknown parameters 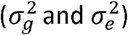 can be efficiently estimated using methods such as Restricted Maximum Likelihood (REML) (Lippert et al. 2011; Yang et al. 2011a). Residual structure in *G* can inflate the genome-wide test statistics, a phenomenon usually summarised by the genomic inflation factor *λ*_GC_, defined as the ratio of the median observed chi-square 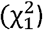 statistic to its null expectation (Devlin and Roeder 1999).

We denote the eigen decomposition of GRM to be *G* = *USU*^*T*^, where *U* is an orthogonal matrix, whose columns are the eigenvectors of *G*, and *S* is a diagonal matrix containing the corresponding eigenvalues.

Substituting this into *Var*(*Y*), we obtain:

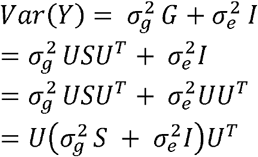

Next, we define a transformation matrix 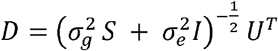.

Subsequently, the transformed variance of *Y* becomes:

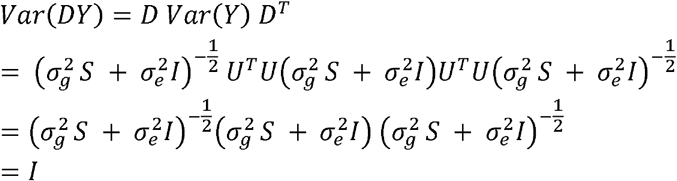

The last step holds because 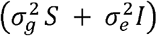is a diagonal matrix.

Now, we apply this transformation to both the phenotype *Y* (with *n* samples) and the genotype *X*(with n samples and p variants), defining 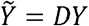 and 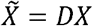. This transformation ensures that the covariance matrix of 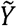 is the identity matrix, i.e., 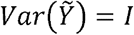.

So, we apply the spectral transformation to the MOKA test to the transformed data, we compute the test statistic:

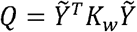

Where 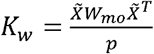 is the kernel matrix and is *W*_*mo*_ a weight matrix that reflects the multi-omics weight specific contribution of each variant and used for MOKA. Spectral decomposition is available as a default functionality in MOKA which can be toggled on or off on the configuration file of the pipeline (see Table 1).

### 2.6. Genomic inflation factor (λGC) calculation

Following Devlin and Roeder (1999), we first convert each gene-level p-value to its equivalent χ^2^ statistic via the inverse cumulative-distribution function (inverse CDF) of a χ^2^ distribution with one degree of freedom. 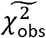 denotes the median of these observed statistics. We then divide this value by the theoretical median of a χ^2^ distribution with one degree of freedom, 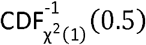:

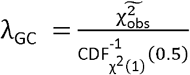

Because the denominator is approximately by theory, λGC ≈ 1 indicates well-calibrated test statistics, whereas λGC > 1.4 signals genomic inflation arising from unmodelled population structure or other confounding effects (Yang et al. 2011b); however they show that even with perfect population structure matching, λGC rises roughly in proportion to polygenicity, so gene - level statistics with many contributing variants can have values well above 1 without implying confounding.

### 2.7. Post-GWAS annotation and Gene Set Enrichment

The output structure is organized into easily navigable folders, including “output_plots” and “result_folder.” KEGG pathway enrichment (snakemake --cores 1 kegg_pathway_analysis) is performed using PathfindR (Ulgen et al. 2019) package, and the Gene Ontology analysis (snakemake --cores 1 go_analysis) is performed with g:Profiler (Reimand et al. 2007) package (Figure 1C).

DisGeNET (Pinero et al. 2017) is a database encompassing 1,134,942 gene-disease associations (GDAs) involving 21,671 genes and 30,170 traits. DisGeNET provides specific summaries for each disease within this platform, detailing gene associations and information on identified significant genes after multiple testing corrections and the proportions associations are in the database (Figure 1E) (snakemake --cores 1 disgenet_annotation_005).

## 3. Results

### 3.1. Configuration of MOKA and multi-omics data bridge

In this demonstration, we used 17-way human and primates accelerated conservation scoring (Siepel et al. 2005; Pollard et al. 2010) as weights (*W*_*mo*_) for each variant. The acceleration measures quantify the extent of human-specific sequence change, such as sites that have diverged more rapidly than expected under neutrality. Negative phyloP (Pollard et al. 2010) scores already encode acceleration as −log_10_ p from a likelihood-ratio test, so negative values (e.g., phyloP ≤ −3, corresponding to p ≤ 10^-3^) signal human-specific evolution and associated disorder. These annotations are of particular interest in schizophrenia, a human-complex disease (Doan et al. 2016; Levchenko et al. 2018), providing a biologically informed basis for SNP aggregation.

In the neural-network weights’ data bridge, weights (*W*_*mo*_) were obtained by taking the element-wise sum of the variant-specific encoder and decoder weight matrices from a variational auto-encoder (VAE) (Kingma and Welling 2013) model as previously described by (Enoma D). The network was trained on the input genotype data, so after tuning and convergence the neural network weights learnt in the training process of the latent representations for reconstruction are extracted (Unterthiner et al. 2020; Herrmann et al. 2024).

### 3.2. Association results

Figure 2 presents a Manhattan plot of association results using negative conservation score– based weights, with genes on chromosomes 1–22 plotted against –log_10_. The Bonferroni-corrected significance threshold (p = 2.64e-06) is shown as a horizontal line. Using human accelerated conservation scores as weights, 89 genes were significantly associated with schizophrenia. The top-associated gene, TMEM17, encodes a transmembrane protein involved in ciliogenesis and neural signaling, and has previously been implicated in schizophrenia (Bigdeli et al. 2021).

**Figure 2.**
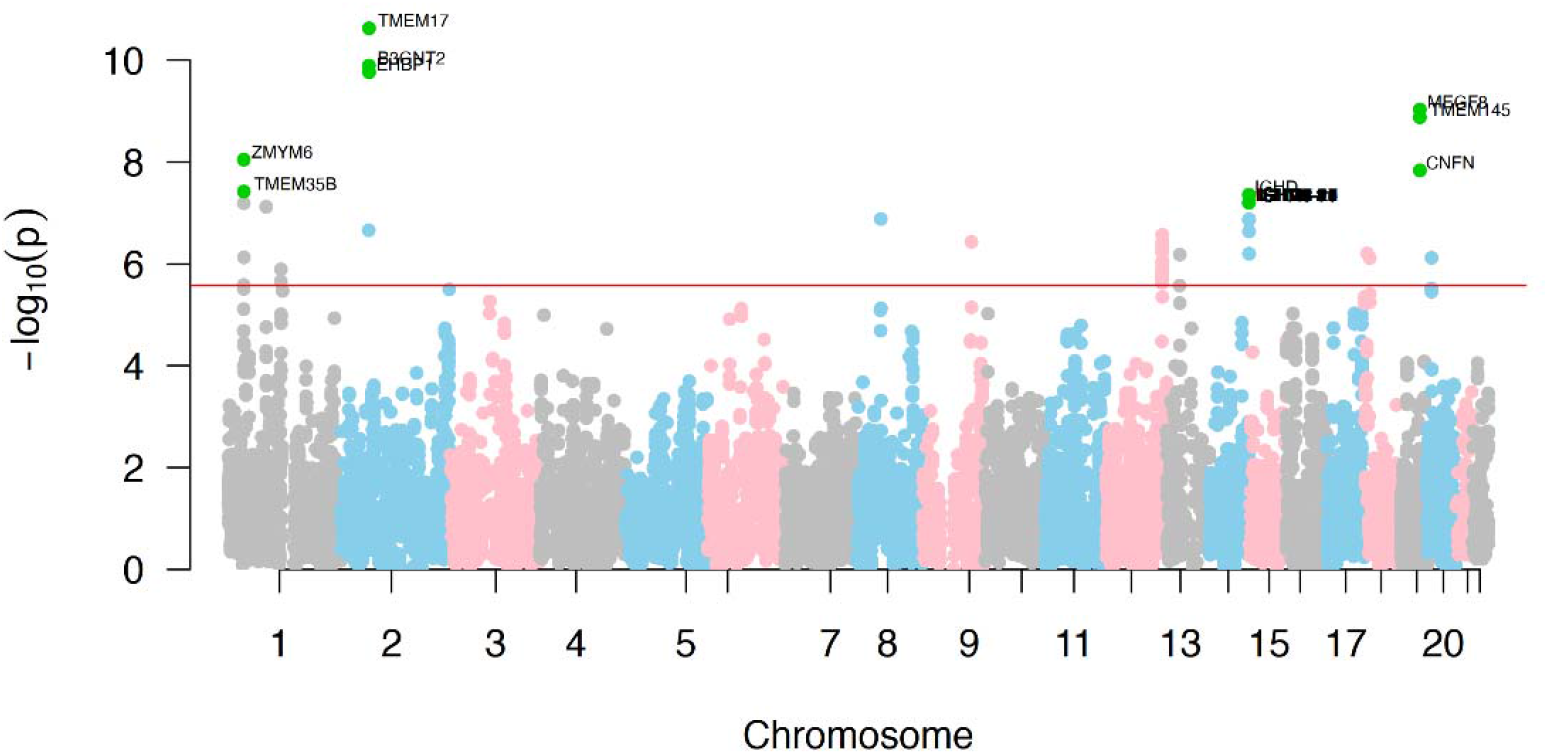
Example Manhattan plot of Association results.

### 3.3. External Database Validation

For validation, we used DisGeNET disease-specific database as a reference database (Schizophrenia in this case). The validation ratio was calculated as the proportion of FDR-significant genes at p-< 0.05 (Benjamini and Hochberg 1995), overlapping with DisGeNET entries, i.e., the number of significant genes found in DisGeNET divided by the total number of significant genes.

Applying the MOKA association test to the schizophrenia GWAS dataset resulted in a 15.7% validation ratio, with 351 associated genes present in DisGeNET. In comparison, SKAT(Wu et al. 2010) yielded a 13.3% validation ratio with 2,530 genes (genomic-inflation factor of λGC ≈ 113), and REGENIE(Mbatchou et al. 2021) yielded 12% with 115 genes validated.

### 3.4. Spectral decomposition and genomic inflation

The MOKA pipeline incorporates spectral decomposition of the variance component to correct inflation in kernel-based tests, improving calibration without sacrificing biological relevance from functional weights (Method). In our schizophrenia GWAS, SKAT(Wu et al. 2010) yielded a highly inflated genomic inflation factor of λGC ≈ 113, indicating substantial confounding and false positives. REGENIE’s whole-genome mixed model reduced this to ≈ 4.7, but lingering stratification remains. Using phyloP acceleration scores in MOKA (MOKA-Ph) reduced SKAT’s inflation to 10.5. Applying MOKA’s spectral correction (MOKA-Ph-COR) further lowered λGC to ≈ 4.3. While still above the ideal benchmark of <1.2, this matches REGENIE’s calibration while retaining mechanistically informed weighting for improved power.

The cohort-wide comparison (Table 2) shows SKAT inflates the test statistics (λGC ≈ 6.9–112.8), while REGENIE reduced this to 3.3–4.7. Incorporating encoder–decoder VAE weights in MOKA (MOKA-ED) reduces the inflation to 4.0–10.1, but remains above REGENIE. The decisive improvement comes from the spectral decomposition of the GRM: MOKA-ED-COR further reduces inflation to 2.5–4.6. In schizophrenia, λGC drops from 113 (SKAT) to 10.1 (MOKA-ED) and 4.6 (MOKA-ED-COR), comparable to REGENIE. These findings demonstrate that MOKA with spectral correction yields well-calibrated, biologically informed association tests, although additional adjustment for population structure may still be beneficial.

**Table 2.**
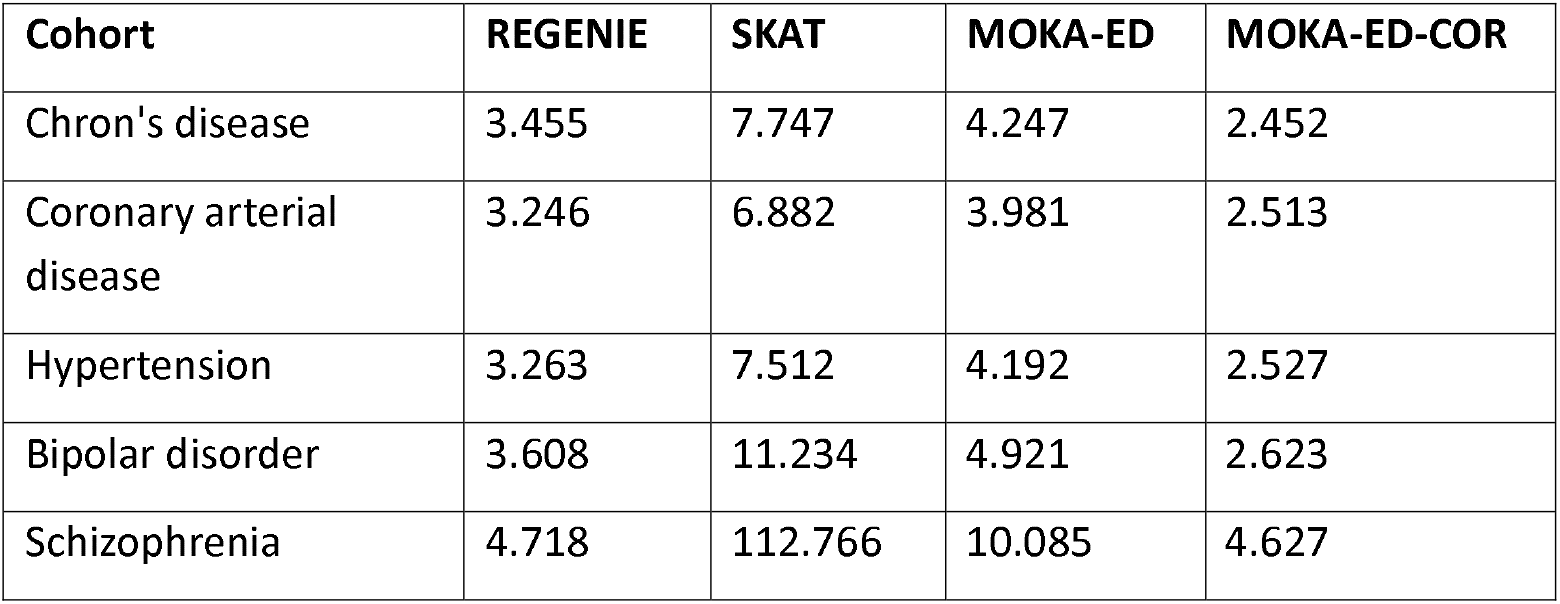
Genomic-inflation factor (λGC) across five cohorts for four association methods (REGENIE(Mbatchou et al. 2021); SKAT(Wu et al. 2010); MOKA-ED = MOKA with encoder– decoder neural-network weights; MOKA-ED-COR MOKA-ED with spectral (GRM) decomposition)

| Cohort | REGENIE | SKAT | MOKA-ED | MOKA-ED-COR |
| --- | --- | --- | --- | --- |
| Chron's disease | 3.455 | 7.747 | 4.247 | 2.452 |
| Coronary arterial disease | 3.246 | 6.882 | 3.981 | 2.513 |
| Hypertension | 3.263 | 7.512 | 4.192 | 2.527 |
| Bipolar disorder | 3.608 | 11.234 | 4.921 | 2.623 |
| Schizophrenia | 4.718 | 112.766 | 10.085 | 4.627 |

### 3.5. Gene Ontology analysis

Gene Ontology analysis in MOKA (Figure 3) of FDR-significant results (p < 0.05) identified top terms for cellular component (cytoplasm, p = 2.0e-31), molecular function (protein binding, p = 5.0e-28), and biological process (anatomical structure development, p = 1.6e-12) for schizophrenia, respectively.

**Figure 3.**
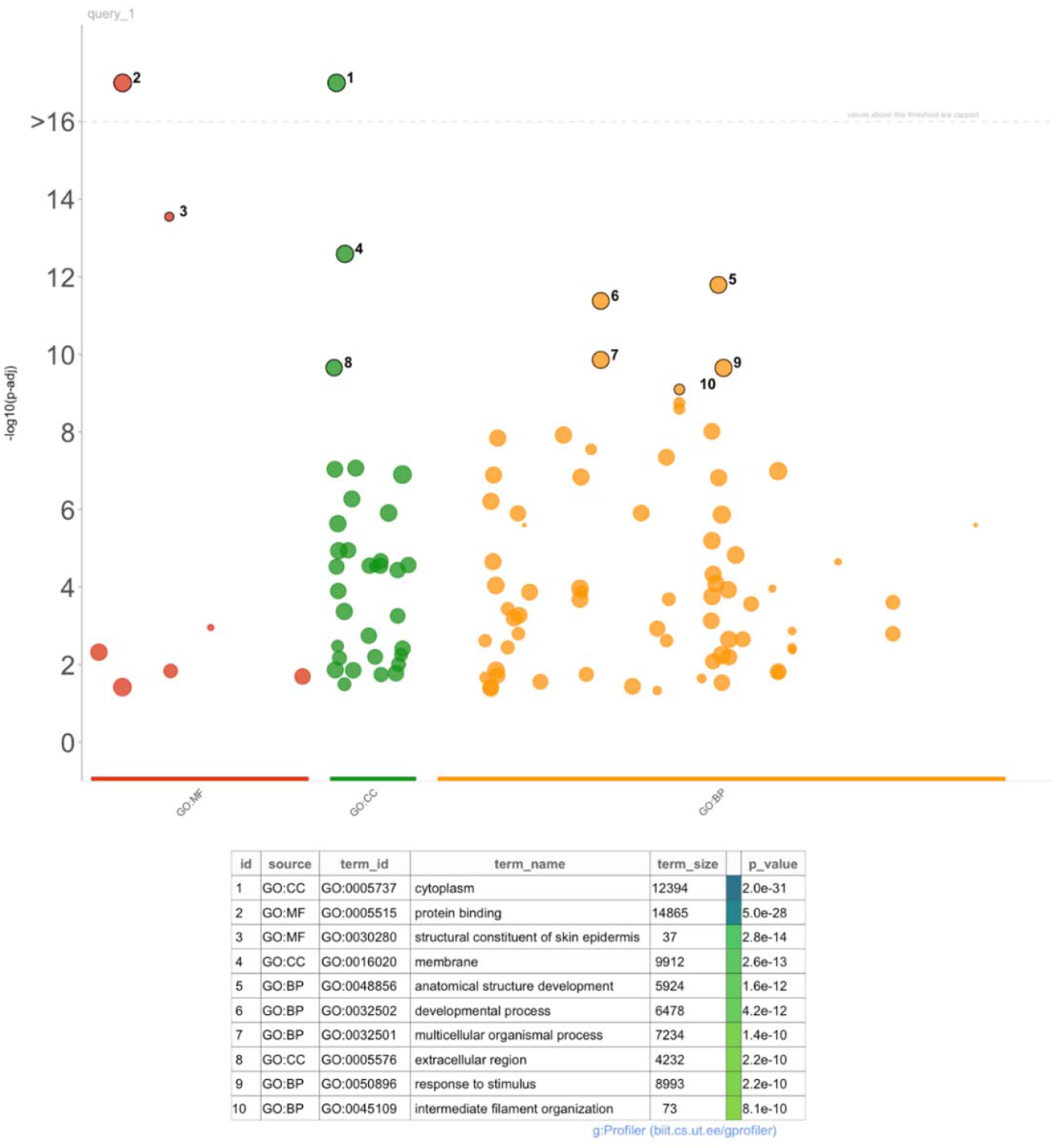
Example Gene Ontology Enrichment results.

### 3.6. KEGG Pathway enrichment

KEGG pathway enrichment analysis in MOKA (Figure 4) of FDR-significant results (p < 0.05) identified several significantly enriched pathways. The top five pathways were Proteasome (p = 1.0e-11), Human Cytomegalovirus Infection (p = 2.0e-10), Nucleocytoplasmic Transport (p = 4.0e-9), prion disease (p = 8.0e-9), and T Cell Receptor Signaling Pathway (p = 1.2e-8).

**Figure 4.**
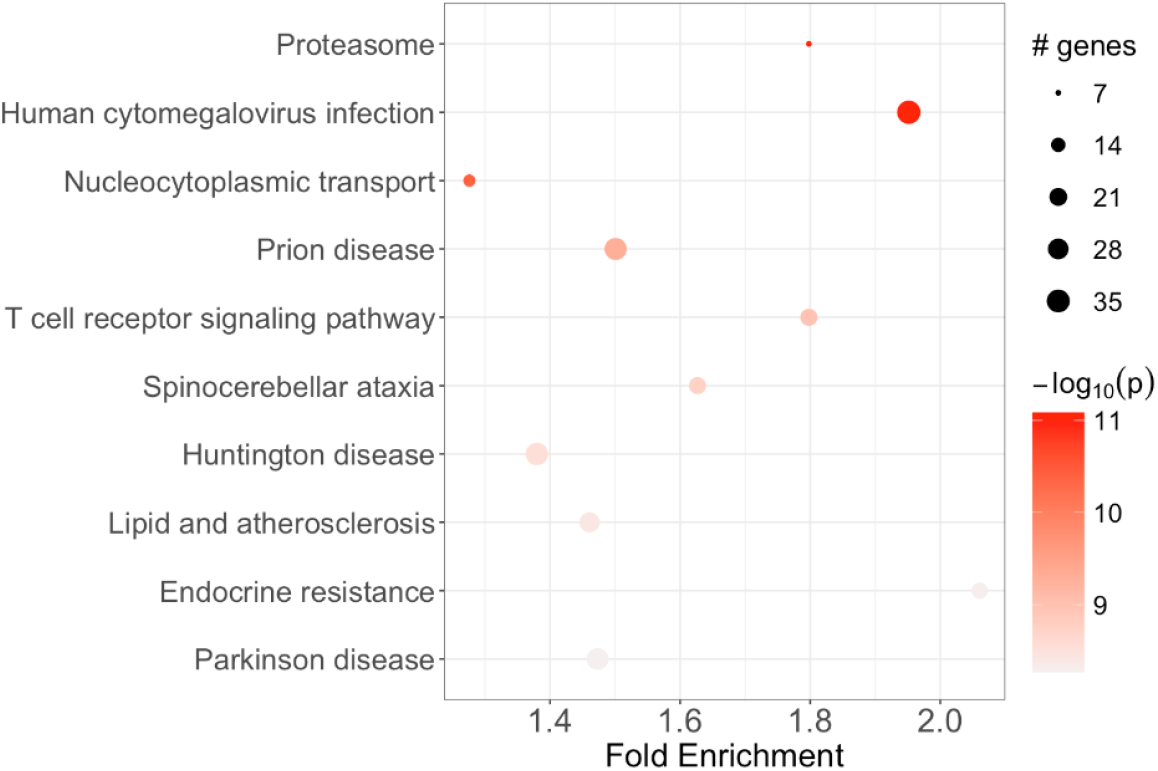
Example KEGG pathway enrichment results

## 4. Conclusion

MOKA is a scalable, automated Snakemake pipeline that enhances GWAS by integrating multi-omics data through SNP-set kernel association tests and applying spectral decomposition to control genomic inflation and population structure. It supports comprehensive post-GWAS analyses and visualization.

In the schizophrenia GWAS, SKAT produced extreme inflation (λGC ≈ 113) with a 13.3% validation rate. MOKA using negative phyloP scores reduced λGC to 10.5 and improved validation to 15.7%. Activating the decorrelation step (“decor-phylo”) further reduced inflation to 4.3. Across WTCCC and schizophrenia datasets, SKAT yielded λGC values between 6.9 and 113, while MOKA with correction reduced this to 2.5–4.6, comparable to REGENIE. Liu et al.(Liu et al. 2013) demonstrated that gene-based tests are susceptible to inflation from gene length and allele-frequency heterogeneity, and that single-SNP genomic control can be overly conservative. MOKA addresses these challenges by providing a robust, flexible, and user-friendly framework for multi-omics integration and kernel-based association analysis, accessible to the broader research community.

## 5. Data and Code availability

The input genotype data is from the Molecular Genetics of Schizophrenia - nonGAIN Sample (MGS_nonGAIN) (dbGaP Study Accession: phs000167.v1.p1) may available at https://www.ncbi.nlm.nih.gov/projects/gap/cgi-bin/study.cgi?study_id=phs000167.v1.p1 and Wellcome Trust Case Control Consortium (WTCCC GWAS)(Wellcome Trust Case Control 2007) are in the public domain. The code of Moka Pipeline is freely available on GitHub under the MIT license and can be found at https://github.com/davidenoma/moka. The Conjoint Health Research Ethics Board (CHREB) at the University of Calgary approved this work with ID REB23-0045_REN. The PhyloP(Pollard et al. 2010) conservation scores for each variant position may be downloaded from the UCSC genome browser(Nassar et al. 2023).

## 6. Acknowledgements

We want to acknowledge group members of the Quan Long Lab (Cumming School of Medicine, University of Calgary) that developed models showing the utility of the data-bridged architecture, which is now automated in the Multi omics kernel-based association testing (MOKA) pipeline to promote ease of use of reproducibility.

